# QFEA - A Method for Assessing the Filtration Efficiency of Face Mask Materials for Early Design Prototypes and Home Mask Makers

**DOI:** 10.1101/2020.12.14.20221937

**Authors:** Eugenia O’Kelly, Anmol Arora, Corinne O’Kelly, Charlotte Pearson, James Ward, P John Clarkson

## Abstract

The COVID-19 pandemic has led to a surge in the design and production of fabric face coverings. There are few published methods which enable mask designers, makers and purchasers to assess the relative filtration ability of mask making materials. Those methods which do exist are prohibitively expensive and difficult to conduct. As a result, mask makers, non-profits, and small-scale designers face difficult decisions when designing face coverings for personal and commercial use. In this paper, we propose a novel method, the Qualitative Filtration Efficiency Assessment (QFEA), for easily and inexpensively comparing the filtration efficiency of common materials. This method provides a highly affordable and readily available way to assess potential mask materials.

## Introduction

The COVID-19 pandemic has led to widespread use of fabric face coverings by the general public. In response to this unprecedented demand, a plethora of new face mask designs are being developed and promoted by individuals and institutions. Unfortunately, most fabric face mask designers, home sewers, and purchasers do not have access to reliable tools by which they can assess the suitability of fabric face mask materials.

It is well established that protective fabric face masks must be constructed of materials with high filtration efficiency. Regrettably, there are few tools available which enable evaluation of the filtration efficiency of filter materials. The tools which are available are prohibitively expensive and is usually inaccessible for home most mask makers and small-scale designers. They are even less accessible to the multitude of individuals purchasing fabric face masks for their families, friends, and institutions. Perhaps at greatest disadvantage are populations with limited access to resources, such as disadvantaged groups and developing countries. Providing designers with easy and inexpensive tools to hone their designs during the earliest stages of the design process is known to be critical to achieving a better design *(1,2)*. Furthermore, prototyping “cheaply and early” has been found to have a strong correlation with positive design outcomes *(3)*.

Although studies exist which assess the filtration efficiency of fabric types, the maker needs to assess the specific filter materials they have access to at home *(4–7)*. Methods which have been proposed for testing filtration efficiency generally require expensive equipment and often utilize particle counters, the supply of which can be expected to be considerably constrained during a pandemic *(4,8,9)*. Early research to explore accessible alternative methods has been conducted; however, these still require equipment and some, albeit limited, expertise which may be unaffordable or unavailable to members of the public or small businesses *(10)*.

This study proposes a novel method, the Qualitative Filtration Efficiency Assessment (QFEA) to: (1) assess whether a filter material has sufficient filtration efficiency to be worth the cost and time incurred to warrant further testing, and (2) compare the filtration effectiveness of multiple filter materials. We present two ways of completing a QFEA. The first method relies on the use of a nebulizer, which can be bought as part of a qualitative fit testing solution, from a medical supply store, or from major retailers such as Amazon. In some areas, the purchase of nebulizers is restricted. The second method builds on the authors’ prior research, utilizing an inexpensive aroma diffuser *(11)*. These are, widely available from major retailers such as Amazon and Target and can be purchased from as little as approximately $18 or £20.

## Methods

### Testing Method

The QFEA method of evaluating filter materials is based on the methods used for qualitative mask fit testing. Qualitative fit testing is a commonly used, OSHA approved method of assessing the fit of a sealing face mask such as an N95 or FFP3 mask.

Qualitative fit testing is based on the knowledge that high filtration materials, such as those used to construct N95 masks, block certain small particles which can generate a taste or smell sensation in the participant. Common solutions used are sodium saccharin which generates a sweet taste, and Bitrex which generates a very bitter taste. If the participant is wearing a well-fit N95 mask, the high filtration efficiency material of the mask will block these particles and the participant will not be able to taste the sweet or bitter solution. If the mask has gaps, not all particles will be filtered as some travel through gaps, thus generating a taste sensation in the participant. Our research team hypothesized that this same basic method could be used to evaluate how effective a material is at blocking small particles. In order to test this, the fit portion of the test which looks for gaps needed to be controlled. When fit was taken out of the equation, we hypothesized that the qualitative fit test would provide an estimate of a material’s filtration ability.

A material with high filtration efficiency, such as an N95 mask, is known to filter out all taste molecules. We hypothesized that the amount of taste present when breathing a sweet or bitter solution through other materials would correlate with the filtration efficiency of those materials.

### Testing Apparatus

Tests were conducted with two qualitative fit testing set-ups: (1) a home-made sodium saccharin formula of 830 mg to 1ml distilled water using an aroma diffuser, and (2) a commercial Bitrex solution with a qualitative test kit nebulizer *(11)*.

An aroma diffuser is likely to be the most cost-effective and easily obtainable QFEA setup. However, those choosing to use commercial qualitative fit testing solution rather than homemade solution may prefer a nebulizer. While homemade qualitative fit testing solution can be made economically, the commercial solution may be expensive and provided in very small quantities not compatible with the aroma diffuser. We thus tested a standard issue medical nebulizer, widely sold for the treatment of health conditions, as an alternative.

Material samples were prepared so that no leaks would be present and all of the participants’ breath would travel through the material. Short, 1” diameter pieces of PVC pipe were used as mouth pieces (see Fig. 1). Potential filtration material samples were used to cover one end of the PVC pipe and secured. Participants were instructed to hold the sample tubes tightly to their mouth, material side out, and breathe normally through the tube. Participants were instructed not to inhale through their nose during the test.

**Fig. 1:**
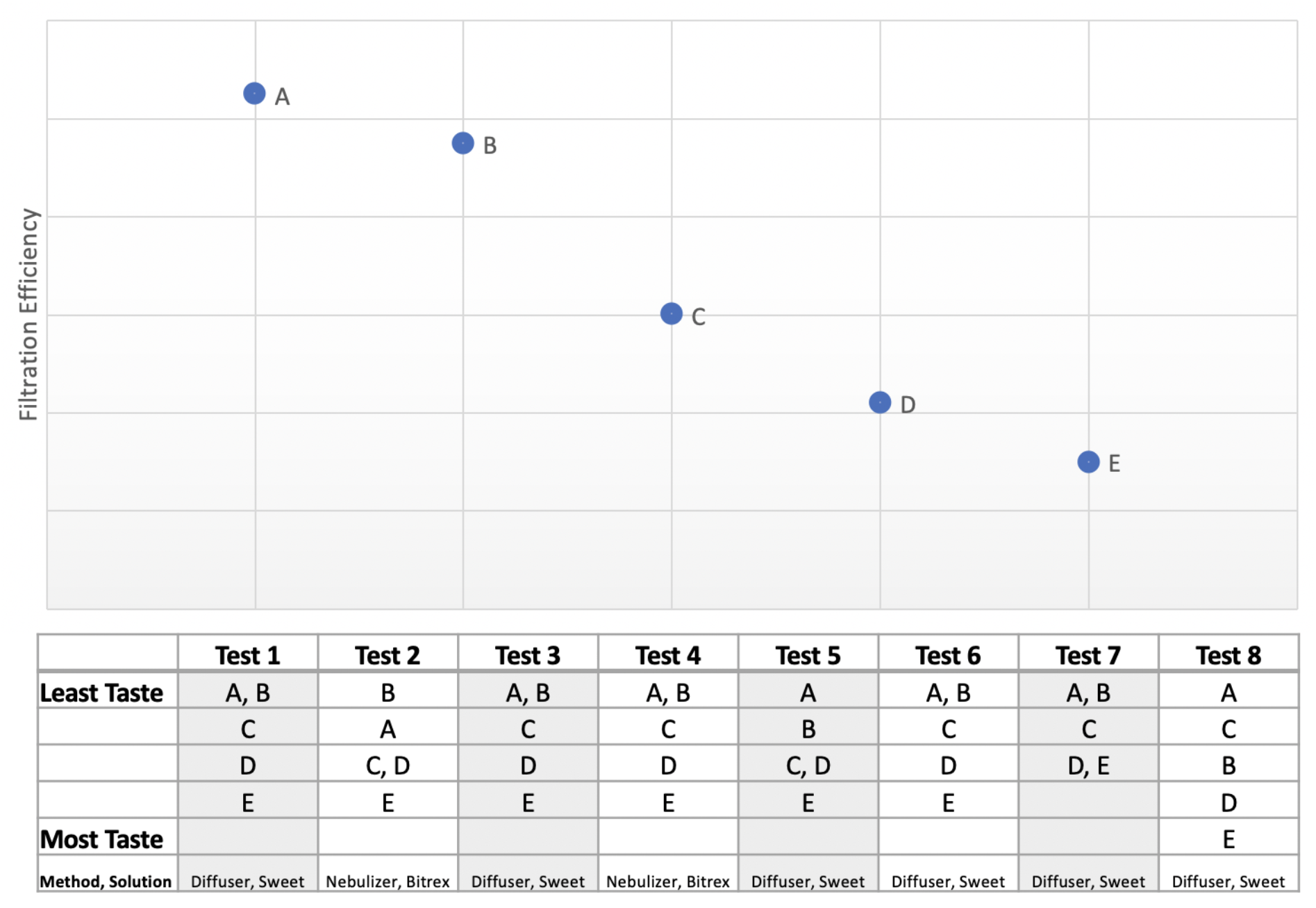
Filtration efficiency of the filter materials compared with the participant’s rankings. If participants were unable to determine which of two filter materials was superior, the two filter materials are placed in the same cell in the feedback table.

Material samples were labeled A-B. Sample A continued filter material from a NIOSH certified N95 mask. Sample B material was taken from a surgical mask. Samples C, D, and E were fabrics previously tested for filtration efficiency *(7)*.

### Testing Procedure

Seven participants of different ages and genders underwent a blind validation study, with two participants taking the study with both sweet and bitter solutions. As when performing a quantitative fit test, the first step of the test is to ensure the participant can taste the solution used. Some participants are known to be unable to taste some solutions, so this step ensures the participant will be sensitive to the solution used *(12)*. Indeed, one participant was unable to taste either the bitter or sweet solutions.

An empty sample tube, labeled X, was provided for this purpose. Participants were instructed to hold the sample tightly to their mouth and inhale the testing solution. If participants could not taste the bitter or sweet solution, the test was discontinued. If participants could taste the solution, we informed them to remember that amount of sensation as a baseline for “Very Strong Taste”.

Participants were then instructed to breathe through samples A through E out of order. In each case, participants were instructed to hold the sample, material side out, closely to their mouth and inhale only through their mouth. If there was a risk a participant might identify samples visually, he or she was blindfolded. After at least 30 seconds with the aroma diffuser or 15 squeezes of the nebulizer, participants were asked to categorize the amount of taste they received as: No Taste; Barely Taste; Some Taste; or Strong Taste. Participants were also instructed to rank-order the samples from the strongest taste to least strong of taste. As participants continued through the samples, they were invited to return to samples they had previously tested to ensure they were confident in their ranking.

The results were then analyzed by the study designer, who compared the participants’ rankings with the known performance of the filter material.

### Comparing Results with Filtration Efficiency

These results were compared to the filtration efficiency of the filter materials, which had previously been evaluated (7).

## Results

As shown in Fig. 2, the correct order of filtration efficiency was A (should produce least taste) to E (should produce most taste). In seven out of the eight tests, participants correctly identified Sample A as the most effective. In five of these cases, Sample B, the surgical mask, was also ranked as having no or almost no taste. In five out of the eight tests, participants were unable to determine if Sample A or Sample B were most effective and ranked them equally.

All samples were correctly ranked in seven out of eight tests. In the test the participant incorrectly ranked Sample B as more effective than Sample A. In another other test, Sample C was incorrectly ranked as having less taste than Sample B.

All subjects correctly identified Sample E as the least effective.

## Discussion

### Summary of Key Findings

Participants were able to accurately class and rank filter materials in a way which reflected the filtration efficiency of the sample. Critically, users were able to judge between high-performance and low-performance filter materials. They were also able to identify which filter materials would be best for mask design. In most cases, participants were able to rank order both fabric and commercial filtration filter materials correctly. Incorrect rankings did occur; these usually occurred when filter materials of similar performance properties were misordered or when participants did not wait long enough between tests and/or failed to rinse out their mouths between tests.

### Implications of Findings

Participants struggled to identify differences between filter materials which performed at similar levels, such as between the surgical mask and N95 mask fabric. This rendered participants unable to determine which of the two filter materials with high filtration efficiency were superior. Participants were more successful in ranking the lowest-functioning filter materials, with all participants correctly sorting the two filter materials with the lowest filtration efficiency. Overall, this suggests that the method described may offer a crude estimate of whether a filter material offers a high filtration efficiency; however, more robust methods, including quantitative testing, would be required for comparing similar filter materials. The QFEA can be used by designers and makers to eliminate low functioning filter materials and focus only on higher functioning filter materials.

### Study Limitations

Not every participant was able to taste each solution. In fact, one participant was unable to taste either the sweet or bitter solution. Other participants had a diminished ability to taste one or both of the solutions. This issue has been well identified in qualitative fit testing upon which this evaluation method is based. As with any qualitative fit tests, the results are subjective and depend on the sensitivity of the tester.

When conducting back-to-back tests, ‘aftertaste’ from the prior sample was found to sometimes affect the evaluation of the succeeding sample. Buildup of taste continued to be a problem, with participants tending to report mild taste sensations for high-performance filter materials if they were presented later in the test or chose not to use the provided water to rinse their mouth and clear existing taste. To help mediate this, participants were asked to take breaks between samples and/or drink water to clear their palate. This was believed to be the case during Test 8, where the participant did not take the opportunity to drink water or take breaks between tests.

## Conclusion

The dearth of reliable face masks for the general public during the COVID-19 pandemic has led to the use and creation of fabric face coverings across the world. The first hurdle in designing or purchasing a face mask is to choose a filter material with the highest filtration as possible while also affording the wearer the highest level of comfort and breathability. The lack of affordable methods to even grossly ascertain the filtration efficiency of filter materials has led to an unfathomable number of masks being constructed, marketed, purchased and used worldwide that have unknown filtration capacity.

A possible approach to this dilemma is adopting the Qualitative Filtration Efficiency Assessment (QFEA). Utilizing the QFEA provides an affordable method for anyone contemplating designing, purchasing or wearing a fabric face mask to gain insight into which of the various options available affords the highest filtration.

The Qualitative Filtration Efficiency Assessment (QFEA) has neither the accuracy nor specificity to act as a replacement for laboratory testing of a filter material’s filtration efficiency; however, our evaluation study showed the QFEA was a useful tool in comparing potential filter materials. It allowed participants to identify the most promising filter materials with the highest filtration efficiency.

It is important to note that the methods described in this paper are not intended to act as substitutes for conventional testing. Laboratory testing remains critical for any respiratory protection tool which is to be introduced to the market. The QFEA offers a simple test for the general public to assess the filtrative ability of masks and a complementary tool for early-stage design, which may be used in addition to existing tests.

## Data Availability

All eessential data is contained within the article's body.

